# Risk of new-onset Long Covid following reinfection with SARS-CoV-2: community-based cohort study

**DOI:** 10.1101/2023.04.13.23288522

**Authors:** Matthew L. Bosworth, Boran Shenhuy, A. Sarah Walker, Vahé Nafilyan, Nisreen A. Alwan, Margaret E. O’Hara, Daniel Ayoubkhani

**Affiliations:** Data & Analysis for Social Care and Health Division, Office for National Statistics, Newport, UK; Methodology and Quality Directorate, Office for National Statistics, Newport, UK; National Institute for Health Research Health Protection Research Unit in Healthcare Associated Infections and Antimicrobial Resistance, University of Oxford, Oxford, UK; Nuffield Department of Medicine, University of Oxford, Oxford, UK; Faculty of Public Health, Environment and Society, London School of Hygiene & Tropical Medicine, London, UK; School of Primary Care, Population Sciences and Medical Education, Faculty of Medicine, University of Southampton, UK; NIHR Southampton Biomedical Research Centre, University of Southampton and University Hospital Southampton NHS Foundation Trust, Southampton, UK; NIHR Applied Research Collaboration (ARC) Wessex, Southampton, UK; Long Covid Support, Registered Charity, London, UK; Leicester Real World Evidence Unit, Diabetes Research Centre, University of Leicester, Leicester, UK

**Keywords:** COVID-19, Coronavirus, Long Covid, Post-COVID Condition, Reinfection

## Abstract

**Background:** Little is known about the risk of Long Covid following reinfection with SARS-CoV-2. We estimated the likelihood of new-onset, self-reported Long Covid after a second SARS-CoV-2 infection, and compared to a first infection.

**Methods:** We included UK COVID-19 Infection Survey participants who tested positive for SARS-CoV-2 between 1 November 2021 and 8 October 2022. The primary outcome was self-reported Long Covid 12 to 20 weeks after each infection. Separate analyses were performed for those <16 years and ≥16 years. We estimated adjusted odds ratios (aORs) for new-onset Long Covid using logistic regression, comparing second to first infections, controlling for socio-demographic characteristics and calendar date of infection, plus vaccination status in those ≥16 years.

**Results:** Overall, Long Covid was reported by those ≥16 years after 4.0% and 2.4% of first and second infections, respectively; the corresponding estimates among those <16 years were 1.0% and 0.6%. The aOR for Long Covid after second compared to first infections was 0.72 (95% confidence interval: 0.63–0.81) for those ≥16 years and 0.93 (0.57–1.53) for those <16 years.

**Conclusions:** The risk of new-onset Long Covid after a second SARS-CoV-2 infection is lower than that after a first infection for those ≥16 years, though there is no evidence of a difference in risk for those <16 years. However, there remains some risk of new-onset Long Covid after a second infection, with around 1 in 40 of those ≥16 years and 1 in 165 of those <16 years reporting Long Covid after a second infection.

## Introduction

Long Covid describes symptoms such as fatigue, breathlessness, pain, and cognitive impairment that persist for months or years after a SARS-CoV-2 infection. As of 2 January 2023, 2 million people in the United Kingdom (3.1% of the population) were estimated to be experiencing Long Covid, with 1.5 million of these reporting limitations to their daily activities [1]. SARS-CoV-2 reinfection rates increased rapidly following the emergence of the Omicron variant and remain high. More than 90% of reinfections occurred during the period when the Omicron variants were dominant; as of 23 November 2022, the estimated rate of reinfection was 40.6 per 100,000 participant days at risk, compared with 11.5 as of 13 December 2021 (before Omicron was the dominant variant) [2]. However, there is limited evidence regarding the risk of new-onset Long Covid following SARS-CoV-2 reinfection.

Descriptive data from a survey administered by Long Covid patient support groups in the UK suggest that most respondents with Long Covid (89%) developed it after their first SARS-CoV-2 infection [3]. However, this finding is not generalisable to the whole population as the data were collected from social media support groups for people with Long Covid (i.e., a highly self-selecting group). Another study using data from electronic health records suggests that SARS-CoV-2 reinfection increases the risk of post-acute sequelae such as death and organ-specific impairment up to six months post-infection [4]. However, the study sample of US military veterans is unlikely to be representative of the broader population, and the study did not assess common Long Covid symptoms.

We therefore investigated the risk of new-onset Long Covid following a second SARS-CoV-2 infection and how this compares with first infections, using data from a large community-based sample selected at random from the UK population.

## Methods

### Study data and design

The main data source for this analysis was the UK COVID-19 Infection Survey (CIS, ISRCTN21086382, https://www.ndm.ox.ac.uk/COVID-19/COVID-19-infection-survey/protocol-and-information-sheets), run by the Office for National Statistics (ONS) and comprising a sample of over half a million participants randomly selected from the UK community population (excluding communal establishments such as hospitals, care homes, halls of residence, and prisons).

Ethical approval was obtained from the South Central Berkshire B Research Ethics Committee (20/SC/0195). At enrolment, participants aged ≥16 years provided written consent, including for optional weekly follow-up assessments for one month followed by at least 12 monthly assessments for the majority of participants. Parents and carers provided consent on behalf of those aged 2-15 years, while those aged 10-15 years also provided written assent.

At each follow-up assessment, all participants answered a survey questionnaire including questions on confirmed/suspected SARS-CoV-2 infections and Long Covid symptoms, and provided a nose-and-throat swab for polymerase chain reaction (PCR) testing.

CIS data for participants in England were linked to Pillar 1 (swab testing for SARS-CoV-2 in UK Health Security Agency laboratories and NHS hospitals for those with a clinical need, and health and care workers) and Pillar 2 (swab testing for SARS-CoV-2 in the wider population, through commercial partnerships, either processed in a laboratory or more rapidly via lateral flow device tests) SARS-CoV-2 test results [5]. To classify COVID-19 vaccination status and timing for participants in England, we used CIS responses linked to National Immunisation Management System (NIMS) records, with the latter being used when data conflicted. Vaccination information for participants in Wales, Scotland, and Northern Ireland was obtained from CIS responses alone.

We included CIS participants who tested positive for SARS-CoV-2 using PCR tests obtained from national testing programmes (participants in England) or during CIS follow-up (all participants), and self-reported positive swab tests (PCR or lateral flow tests) taken outside of the CIS. Among these participants, we identified first and second infections meeting the inclusion criteria **(Figure 1)**; for more details see the Supplementary Methods.

**Figure 1:**
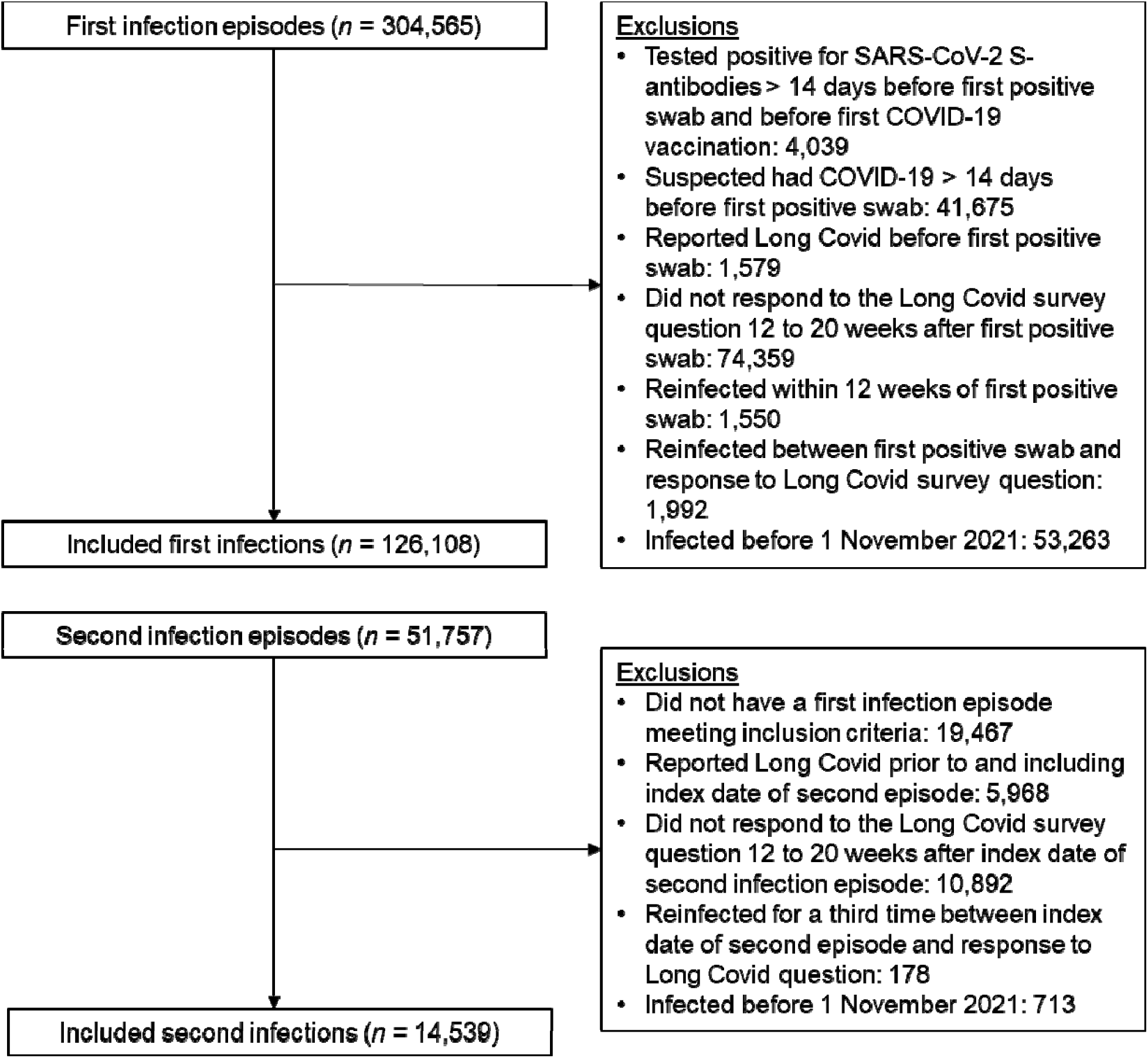
Study participant flow diagram

We then excluded any infections occurring before 1 November 2021. This date was chosen to ensure a reasonable degree of overlap in the calendar date of infection between first and second infection episodes; the fifth percentile of the calendar date distribution was 6 December 2020 for first infections but 13 November 2021 for second infections among those ≥16 years; and 10 December 2020 and 20 October 2021, respectively, among those <16 years **(Supplementary Figure 1)**.

### Exposure

The exposure was a second versus a first SARS-CoV-2 infection, defined by adapting previous methods used for producing official statistics relating to SARS-CoV-2 surveillance in the UK [6, 7]. For more information, see the Supplementary Methods.

### Outcome

The primary outcome was new-onset Long Covid of any severity according to the survey question: “Would you describe yourself as having Long Covid, that is, you are still experiencing symptoms more than 4 weeks after you first had COVID-19, that are not explained by something else?” Participants who responded positively to this question were then also asked about the extent to which their symptoms limited their ability to undertake daily activities (a lot, a little, or not at all), and the presence or absence of 21 individual symptoms attributed to Long Covid (the most commonly reported when the survey question was developed [8-10]). The secondary outcome was activity limiting Long Covid (no Long Covid or Long Covid without activity limitation versus activity limited a little or a lot by Long Covid).

We considered participants’ first response to these questions 12 to 20 weeks after the date of the first positive swab in each infection episode (the index date).

### Covariates

Covariates included socio-demographic characteristics (age, sex, white or non-white ethnicity, area deprivation quintile group, and self-reported pre-existing health conditions), vaccination status, mode of response to the survey at follow-up for Long Covid (digital or face-to-face interview), calendar date of infection (to account for changes in dominant SARS-CoV-2 variant in circulation and other temporal effects), and the number of days from the index date for each infection episode to follow-up for Long Covid.

### Statistical methods

Separate analyses were conducted for those ≥16 years and <16 years. We compared study participants’ socio-demographic characteristics at the first and second infection using means for continuous variables and proportions for categorical variables, with absolute standardized differences ≥10% indicating a large imbalance between infection episodes [11].

We calculated the crude percentage of participants reporting Long Covid 12 to 20 weeks after each infection episode to estimate the absolute risk of new-onset Long Covid. We also calculated the prevalence of a range of Long Covid symptoms as the percentage of those ≥16 years who reported having Long Covid after each infection. This was not possible for participants <16 years due to small sample sizes.

Adjusted odds ratios (aORs) for Long Covid 12 to 20 weeks post-infection were estimated from binary logistic regression models, comparing second infection episodes to first infection episodes (reference group). For those ≥16 years, models were adjusted for all the covariates outlined above. The models for those <16 years were adjusted for age, sex, calendar date of infection, and the number of days from the index date to Long Covid follow-up due to an insufficient number of events for some levels of the other covariates. We did not adjust for COVID-19 vaccination status in those <16 years because of the high correlation with age and underlying health status; children aged <5 years are not eligible for vaccination in the UK, and uptake has been low among those aged 5 to 11 years (just 5.2% of the population of England in this age group had received two doses of a COVID-19 vaccine by 8 October 2022 [12]). All variables were defined at the index date of each infection episode except mode of response, which was defined at the date of the response to the Long Covid question.

Continuous variables (age, follow-up time, and calendar date of infection) were modelled as restricted cubic splines, with boundary knots at the 10^th^ and 90^th^ percentiles and an internal knot at the median of the distributions. We tested one to five knots and selected one internal knot as this minimised the Bayesian Information Criterion for the models.

As it is possible that the impact of reinfection on the development of new-onset Long Covid varies across different sub-populations, for the primary outcome, we used likelihood ratio tests to test for effect modification of the association between reinfection and new-onset Long Covid, by interacting reinfection with each of the covariates included in the models.

All statistical analyses were performed using R version 3.6 software.

## Results

### Description of the study sample

After applying the study inclusion and exclusion criteria **(Figure 1)**, the analysis included 126,108 first infections (110,844 in those ≥16 years, 15,264 in those <16 years) and 14,539 second infections (11,244 ≥16 years, 3,295 <16 years) occurring between 1 November 2021 and 8 October 2022 **(Table 1)**. Median follow-up time from the start of infection to Long Covid response was 102 days (IQR: 92–112) for those ≥16 years and 101 days (92–111) for those <16 years.

**Table 1.**
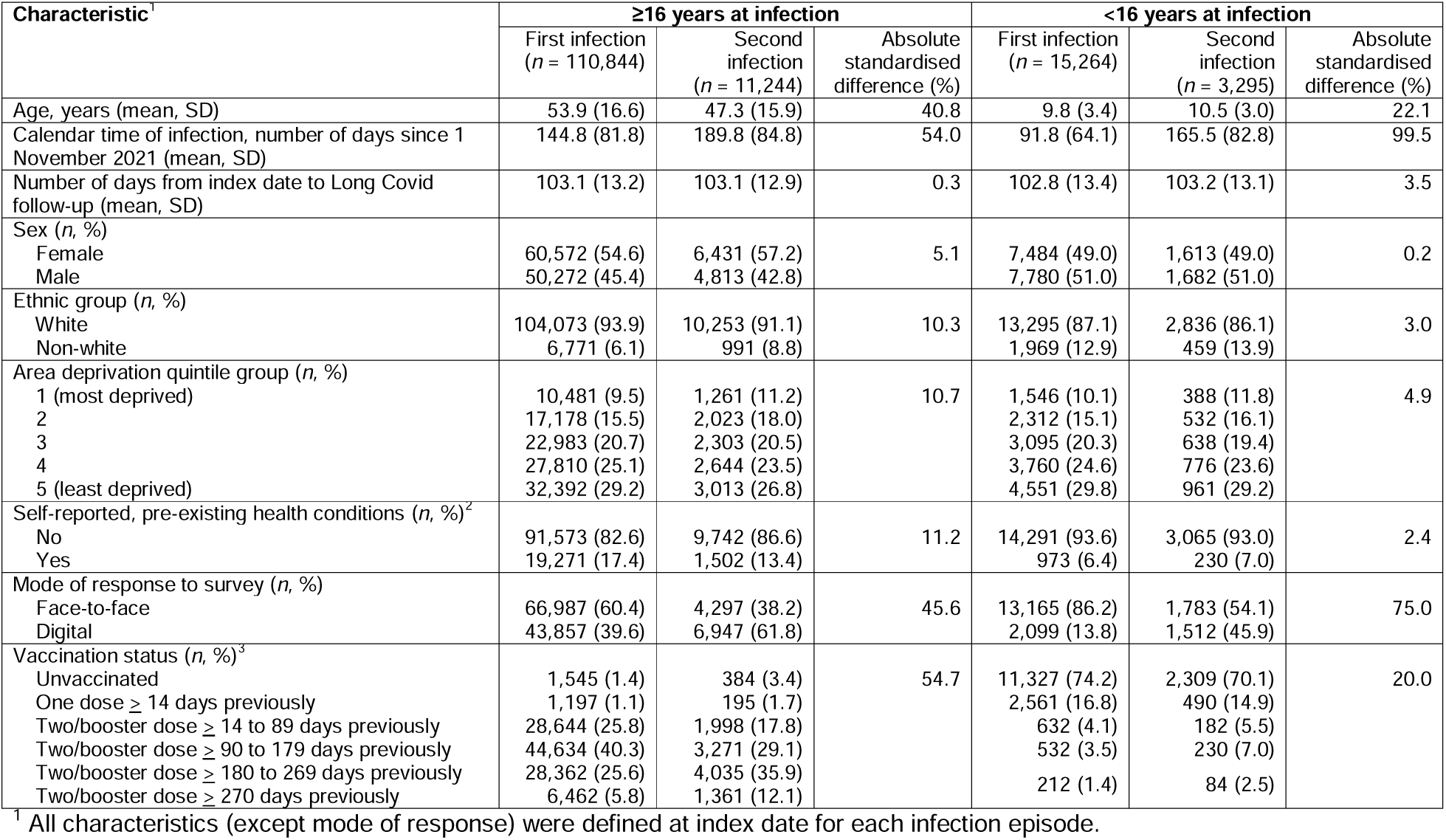
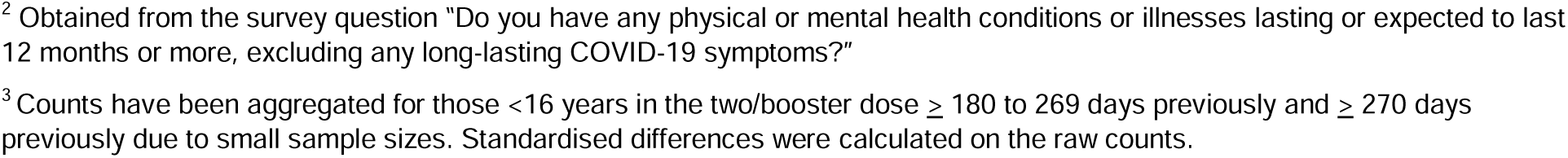
Characteristics of study participants

40.3% of those ≥16 years in the first infection episode group had received two or more doses of a COVID-19 vaccine 90 to 179 days before infection. In the second infection episode group, 35.9% had received at least two doses of a COVID-19 vaccine 180-269 days before infection. Most of those <16 years were unvaccinated in both the first (74.2%) and second (70.1%) infection episode groups.

Among those ≥16 years, the mean age was higher for first infection episodes (53.9 years, SD: 16.6 years) than second infection episodes (47.3 years, SD: 15.9 years) and a larger percentage reported having a pre-existing health condition at the first infection episode (17.4%) than the second infection episode (13.4%).

### Long Covid in those ≥16 years

Long Covid of any severity was reported by 4,381 of those ≥16 years after a first infection (prevalence 4.0%; 95% CI 3.8%–4.1%) and 274 (2.4%; 2.2%–2.7%) following a second infection. Activity limiting Long Covid was reported by 3,103 of those ≥16 years (2.8%; 2.7%–2.9%) after a first infection, compared with 180 (1.6%; 1.4%–1.9%) after a second infection.

The most common symptoms among those ≥16 years with Long Covid were fatigue (61.6% after a first infection, 57.7% after a second infection); shortness of breath (33.7% and 30.7%, respectively); muscle ache (26.7% and 28.5%, respectively), and difficulty concentrating (26.1% and 34.7%, respectively) **(Figure 2)**. The prevalence of neuropsychological symptoms (such as difficulty concentrating, memory loss or confusion, and worry or anxiety) was numerically higher following a second infection. However, small numbers prevented formal statistical testing.

**Figure 2.**
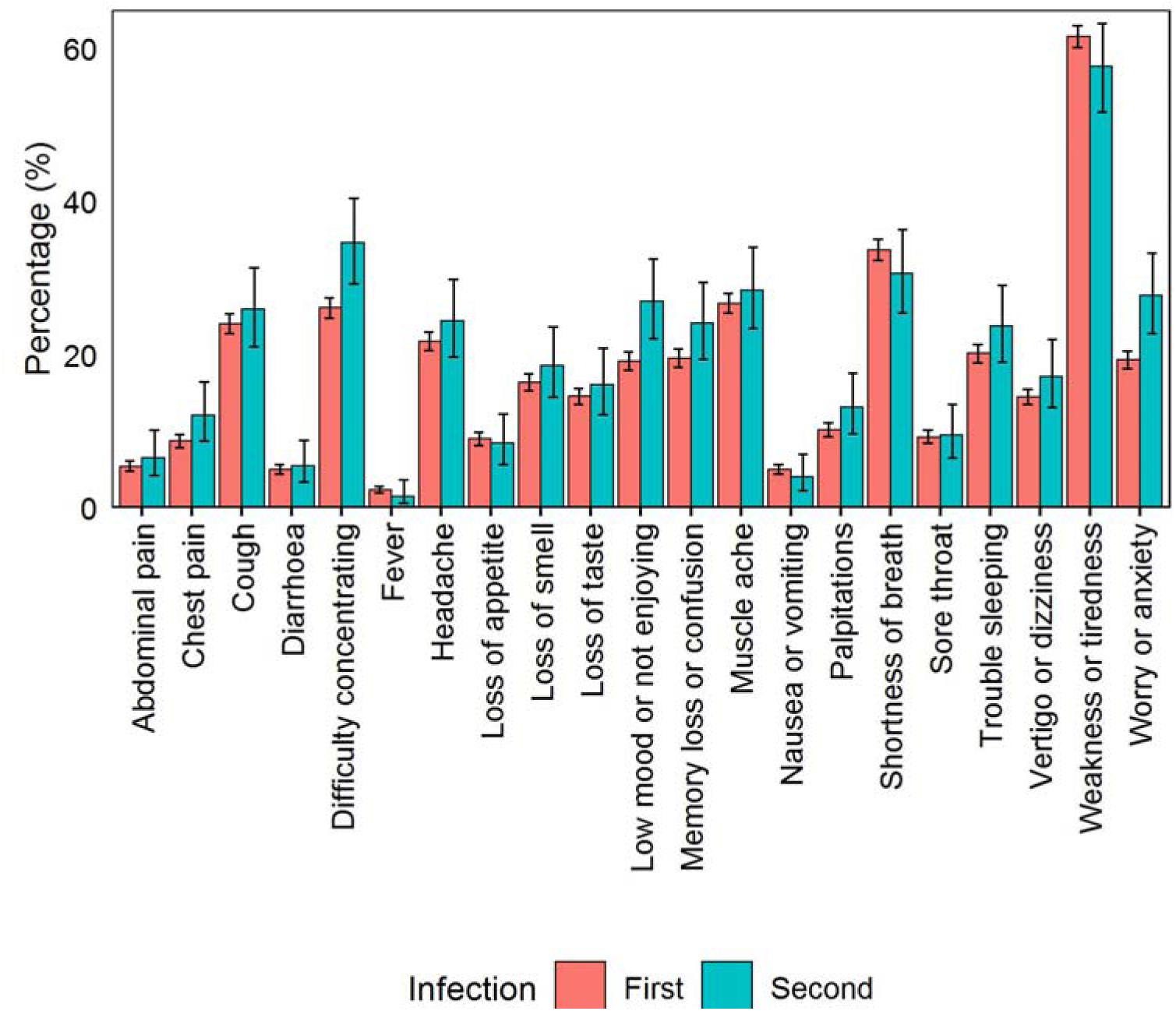
Prevalence of Long Covid symptoms among those ≥16 years who reported having Long Covid after a first or second SARS-CoV-2 infection.

The aOR of reporting Long Covid after a second infection compared to a first infection was 0.72 (95% CI 0.63–0.81) for Long Covid of any severity and 0.66 (0.57–0.77) for activity limiting Long Covid **(Figure 3)**. There was no evidence for effect modification of the association between reinfection and new-onset Long Covid of any severity by age (*p*=0.35), sex (*p*=0.17), ethnicity (*p*=0.98), area deprivation (*p*=0.89), pre-existing health status (*p*=0.14), vaccination status (*p*=0.15), or calendar date of infection (*p*=0.29).

**Figure 3.**
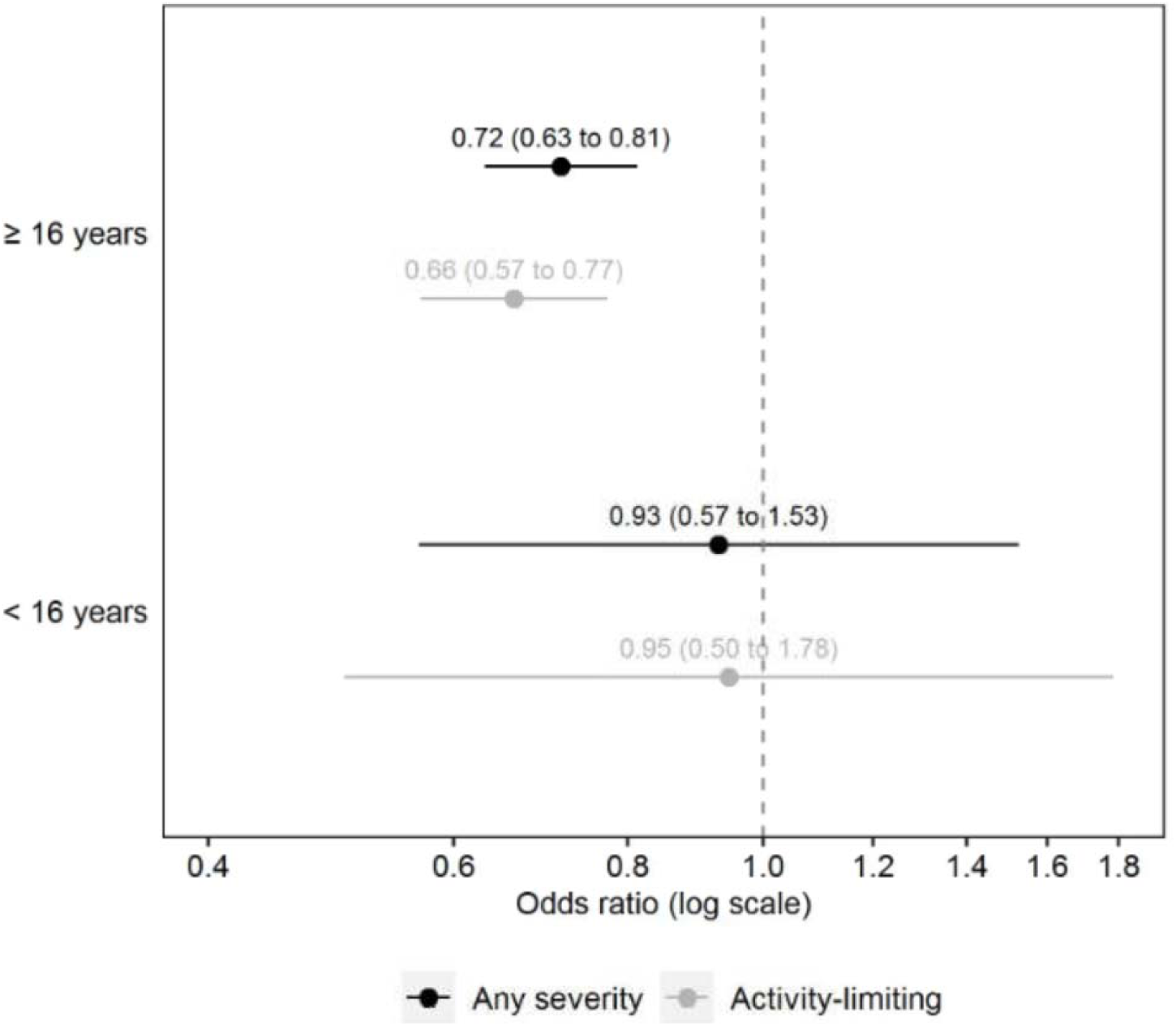
Adjusted odds ratios for Long Covid 12 to 20 weeks after a second SARS-CoV-2 infection compared with a first infection (reference group). Odds ratios for those ≥16 years are adjusted for socio-demographic characteristics (age, sex, white or non-white ethnicity, area deprivation quintile group, and self-reported health status), vaccination status, time from infection to follow-up for Long Covid, calendar date of infection (as a proxy for the dominant SARS-CoV-2 variant in circulation), and mode of response to the survey. Odds ratios for those <16 years are adjusted for age, sex, time from infection to follow-up for Long Covid, and calendar date of infection. Confidence intervals are at the 95% level.

### Long Covid in those <16 years

Long Covid of any severity was reported by 160 of those <16 years after a first infection (1.0%; 0.9%–1.2%) and 20 (0.6%; 0.4%–0.9%) following a second infection. Activity limiting Long Covid was reported by 87 of those <16 years (0.6%; 0.5%–0.7%) after a first infection, compared with 12 (0.4%; 0.2%–0.6%) after a second infection.

The aOR of reporting Long Covid after a second infection compared to a first infection was 0.93 (95% CI 0.57–1.53) for Long Covid of any severity and 0.95 (0.50–1.78) for activity limiting Long Covid **(Figure 3)**. There was no evidence for effect modification of the association between reinfection and new-onset Long Covid of any severity by age (*p*=0.78) or sex (*p*=0.85). The interaction with calendar date of infection was statistically significant (*p*=0.006). However, wide confidence intervals meant there was a high degree of uncertainty around this finding, and the results should be interpreted with caution **(Supplementary Figure 2)**.

## Discussion

### Summary of main findings

Relative to a first SARS-CoV-2 infection, the odds of new-onset Long Covid of any severity or activity limiting Long Covid were 28% and 34% lower, respectively, following a second infection in those ≥16 years, even after adjusting for vaccination status and other potential confounders. This finding may partly be the result of some degree of protection against Long Covid being conferred by prior infection (assuming persistent symptoms were not present after the first infection), coupled with survivorship effects. That is, people with a greater predisposition to Long Covid (for example, females or those with certain underlying health conditions [13]) experiencing persistent symptoms following a first infection, and therefore not being in the sample eligible to experience new-onset Long Covid following a second infection.

In those <16 years, the crude prevalence of new-onset Long Covid was lower following a second infection compared with a first infection, but this difference was not statistically significant after controlling for confounders. However, confidence intervals were wide, reflecting the smaller sample, and compatible with similar reductions to those seen in those ≥16 years.

### Comparison with other studies

Research into the risk of Long Covid following reinfection with SARS-CoV-2 is scarce. Our findings are consistent with descriptive data from self-selecting respondents collected by Long Covid patient support groups, which suggest that the majority of respondents who have Long Covid developed it after their first infection [3]. However, most participants were unvaccinated when they were first infected, and being vaccinated is associated with a reduced risk of developing Long Covid following SARS-CoV-2 infection [14-16]. In addition, reinfections became more common following the emergence of the Omicron variant [2], and the risk of Long Covid has previously been shown to be lower for infections compatible with the Omicron variants compared with the Delta variant [17, 18]. Importantly, our analysis of a randomly selected community-based cohort shows that the risk of new-onset Long Covid in those ≥16 years is lower following a second infection even after adjusting for vaccination status and calendar date of infection (as a proxy for the dominant SARS-CoV-2 variant in circulation at any given time). However, it is important to note that the population prevalence of Long Covid in the UK has remained relatively stable since the emergence of the Omicron variant due to higher infection rates compared with earlier periods in the pandemic [1].

Although the risk of new-onset Long Covid in those ≥16 years was lower after a second SARS-CoV-2 infection than a first infection, the absolute risk is not negligible; 2.4%, that is around one in 40, of those ≥16 years who did not report Long Covid after their first infection went on to do so after a second infection. Other evidence suggests that SARS-CoV-2 reinfection increases risk of post-acute, multi-organ sequelae up to six months after reinfection, compared with a single infection [4]. Our study extends these findings by examining the relationship between reinfection and common Long Covid symptoms. We found that most symptoms reported by those ≥16 years with new-onset Long Covid after a second infection were reported at similar levels of prevalence by participants with Long Covid after a first infection. However, there was some descriptive evidence that the prevalence of neuropsychological symptoms (such as difficulty concentrating, memory loss or confusion, and worry or anxiety) was higher among participants reporting new-onset Long Covid after a second infection, compared with those who reported it after a first infection.

The aim of our study was to estimate the risk of new-onset Long Covid after reinfection, rather than the incremental risk conferred by reinfection in addition to that from the primary infection. Several studies have shown that previous infection with SARS-CoV-2 is associated with reduced risk of severe disease and hospital admission following reinfection, with the strongest association in those with hybrid immunity from vaccination and prior infection [19-21]. Since the pathophysiology of Long Covid is poorly understood [22], future research should investigate the biological mechanisms underlying the association between previous immunity and the reduction in risk of developing Long Covid observed in this study. This could improve understanding of the pathogenesis of Long Covid and potentially improve therapeutics.

### Strengths and limitations

The main strength of the analysis is the use of data from CIS, comprising approximately half a million people randomly sampled from private households to minimise selection bias. CIS participants are routinely tested for SARS-CoV-2, so our study sample included initially asymptomatic as well symptomatic infections. We adjusted for a wide range of factors that may be related to both the risk of reinfection [2] and developing Long Covid [13, 15]. However, the observational nature of the study means that unmeasured confounding may remain, and thus causality cannot be inferred. In particular, we were only able to adjust for age, sex, calendar date, and follow-up time in the analysis of those <16 years due to limited sample sizes.

The routine testing in CIS also means that we can more completely ascertain infection history compared with using results from national testing programmes or self-report alone. We exploited multiple sources of information, including genetic sequencing, S-gene target positivity, and Ct values to distinguish as much as possible between persistent PCR positivity and new infections. However, one limitation is that inevitably some short infections and/or reinfections may have been missed.

We excluded participants who were reinfected less than 12 weeks after their first infection or before they had responded to the Long Covid question 12 to 20 weeks after their first infection. Although only a small number of participants (*n*=3,542, 1.2% of the original sample of first infections) were excluded for this reason, this could introduce bias if a shorter duration of first infection is related to the risk of Long Covid. Consequently, the results may not be generalisable to people who are reinfected with short intervals between their first and second infection.

Another limitation is that Long Covid status was self-reported, so outcome misclassification is possible. Some participants may have been experiencing symptoms because of a health condition unrelated to COVID-19, while others who did have Long Covid may not have described themselves as such (for example, due to the perceived stigma associated with the condition [23]). Conversely, self-recognition of Long Covid (participants’ perception of the change in their own health compared with pre-infection) may be more reliable than electronic health records in some respects, for example due to differences in healthcare-seeking behaviours between socio-demographic groups and Long Covid diagnoses being under-recorded in primary care [24].

This analysis only includes infections occurring between 1 November 2021 and 8 October 2022. The Omicron COVID-19 variant was first identified in the UK on 27 November 2021 [25] and quickly became the main variant in circulation. Most first and second infections in our sample are therefore Omicron infections, and it is unclear whether our findings are representative of infections with other SARS-CoV-2 variants.

## Conclusions

The risk of new-onset Long Covid after a second SARS-CoV-2 infection is lower than that after a first infection for those ≥16 years even after adjusting for vaccination status and variant (using calendar date as a proxy). Although there was no statistical evidence of a difference in risk between first and second infections for those <16 years, there was a large degree of uncertainty around the point estimate, suggesting this finding could be a consequence of lower power in this smaller subgroup. Despite our finding that reinfection carries a lower risk of new-onset Long Covid than a first infection in those ≥16 years, there remains some risk of new-onset Long Covid, following around one in forty second infections among those ≥16 years. Further research is required to understand whether the risk of Long Covid is reduced with each subsequent infection. This is essential to model the expected future burden of Long Covid on the population.

## Data Availability

De-identified study data are available to accredited researchers in the ONS Secure Research Service under part 5, chapter 5 of the Digital Economy Act 2017. For further information about accreditation, contact or visit: https://www.ons.gov.uk/aboutus/whatwedo/statistics/requestingstatistics/approvedresearcherscheme.

## Notes

## Author contributions

MLB, DA, and BS conceptualised and designed the study. MLB and BS prepared the study data and performed the statistical analysis. All authors contributed to interpretation of the results. MLB and DA were responsible for the first draft of the manuscript. All authors contributed to critical revision of the manuscript and approved the final manuscript.

## Patient involvement

MEO and NAA have lived experience of Long Covid.

## Disclaimer

The views expressed are those of the authors and are not necessarily those of the National Health Service, the National Institute for Health Research (NIHR), the Department of Health and Social Care, or the UK Health Security Agency. For the purpose of open access, the authors have applied a Creative Commons Attribution (CC BY) licence to any Author Accepted Manuscript version arising.

## Financial support

The CIS is funded by the Department of Health and Social Care with in-kind support from the Welsh Government, the Department of Health on behalf of the Northern Ireland Government, and the Scottish Government. There was no dedicated funding for this study of CIS data.

*Acknowledgements*

DA is supported by the National Institute for Health Research (NIHR) Applied Research Collaboration East Midlands (ARC EM). ASW is supported by the NIHR Health Protection Research Unit in Healthcare Associated Infections and Antimicrobial Resistance (NIHR200915), a partnership between the UK Health Security Agency (UKHSA) and the University of Oxford. ASW is also supported by the NIHR Oxford Biomedical Research Centre and is an NIHR Senior Investigator. NAA is a co-investigator on the NIHR-funded STIMULATE-ICP and HI-COVE studies and has contributed in an advisory capacity to WHO and the EU Commission’s Expert Panel on effective ways of investing in health meetings in relation to post-COVID-19 condition.

## Supplementary Materials

### Supplementary Methods

#### Inclusion & exclusion criteria

To identify first SARS-CoV-2 infection episodes, we excluded participants who reported suspected COVID-19 or tested positive for S-antibodies (in the study or elsewhere, ignoring blood tests taken after first COVID-19 vaccination) >2 weeks before their first positive swab; reported Long Covid symptoms at any time before their first positive swab; did not respond to the survey question on Long Covid 12 to 20 weeks after their first positive swab; or were reinfected within 12 weeks of their first positive swab or before their first response to the Long Covid question 12 to 20 weeks after their first positive swab (since, if these participants experienced Long Covid, it is uncertain whether their symptoms were attributable to the first or second infection).

To identify second SARS-CoV-2 infection episodes, we excluded participants with a second episode who did not have a first infection episode meeting the above criteria; reported Long Covid prior to (and including) the start of their second episode; did not respond to the Long Covid question 12 to 20 weeks after the start of their second episode; or were reinfected again before their first response to the Long Covid question 12 to 20 weeks after the start of their second episode.

#### Exposure definition

Positive swab test results from any source were grouped into infection episodes to allow for long duration of PCR positivity in some individuals, incorporating information from genetic sequencing, S-gene target positivity and cycle threshold (Ct) values, together with negative PCR test results from CIS only. We defined a new infection episode as a new swab positive occurring >120 days after an index positive with the preceding test being negative, or >90 days with the preceding two consecutive tests being negative (one negative after 20 December 2021 when Omicron variants dominated given higher reinfection rates with Omicron), or >60 days with the three preceding consecutive tests being negative, or after 4 preceding consecutive negative test results at any time.

We further split these infection episodes if they contained multiple sequences from different genetic lineages (e.g., BA.5 and BA.2), or had incompatible S-gene target positivity with Ct<30 (e.g., S-gene positive and S-gene negative, both with Ct<30), or had large decreases in Ct within a set of positive tests grouped together, or low Ct long after the first positive within an episode (both indicative of a new infection rather than ongoing PCR positivity). We also split infection episodes where a new lateral flow device positive was recorded 27 days or more after the start of an infection episode, or 19 days or more after a previous positive PCR or lateral flow test, since this again indicates high viral load and actively replicating virus (more likely associated with a new infection).

**Supplementary Figure 1.**
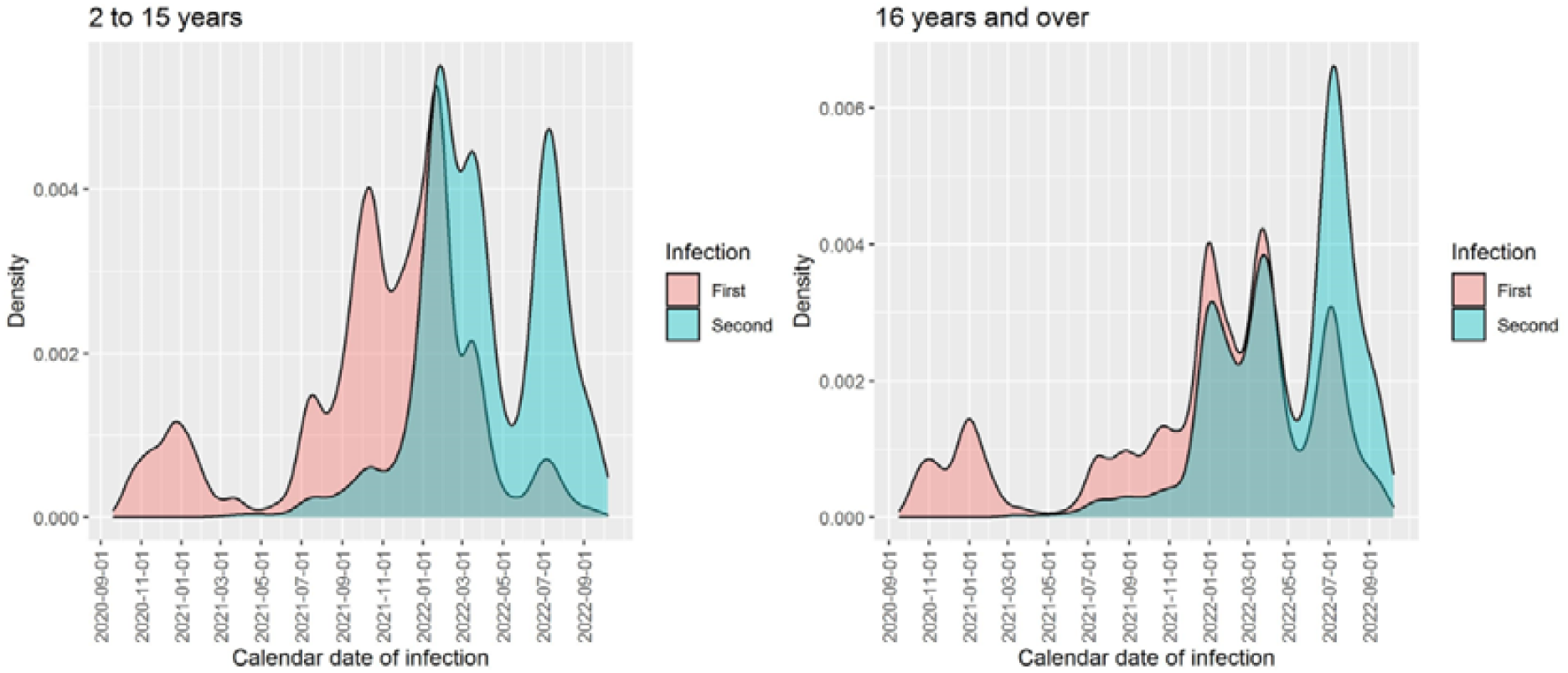
Density plots of calendar date of infection, stratified by infection episode and age group.

**Supplementary Figure 2.**
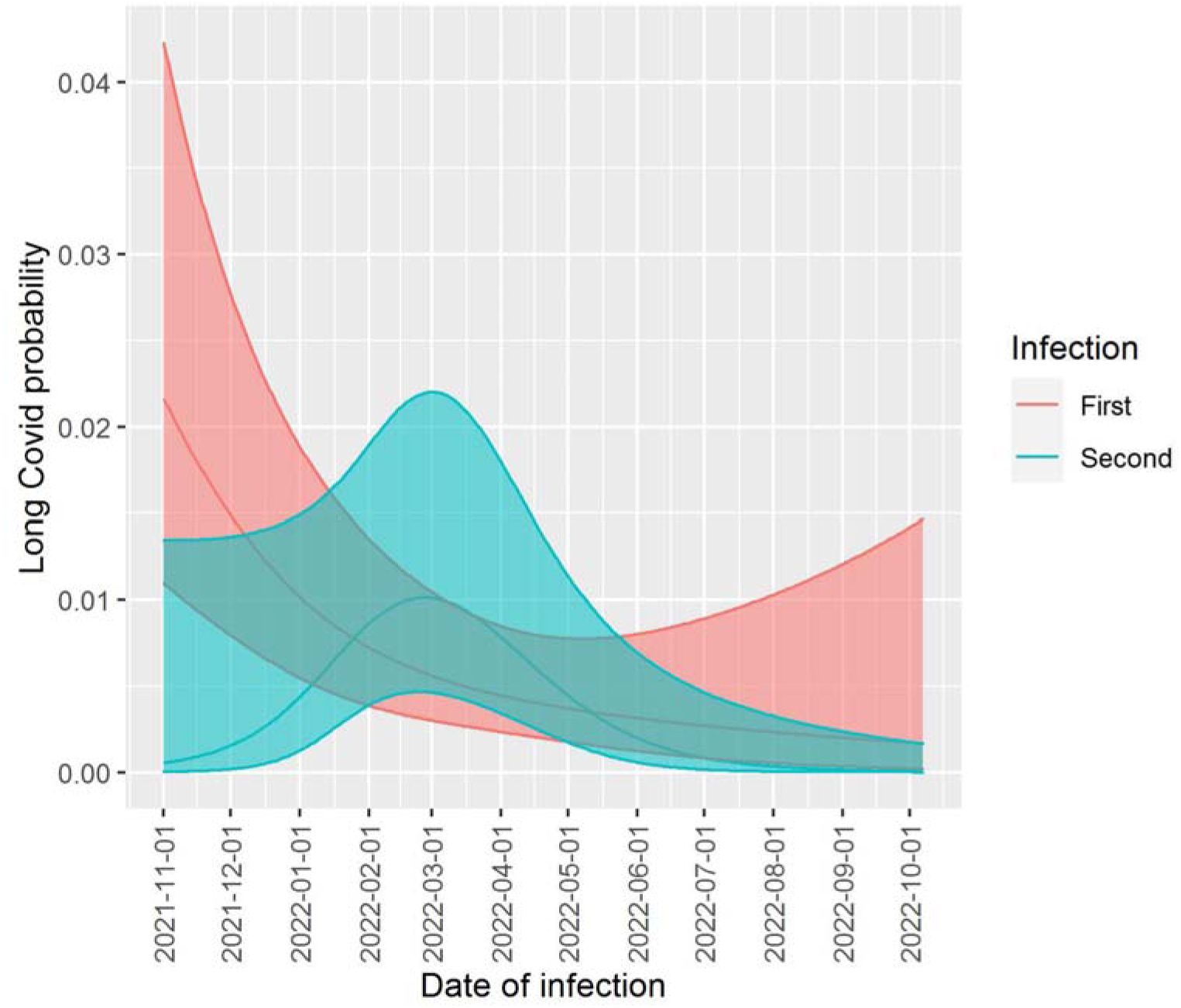
Estimated marginal probability of Long Covid by calendar date of infection in those <16 years.^1^ ^1^Estimates were calculated using the emmeans package, adjusting for age, sex, and time from infection to follow-up for Long Covid. Shaded areas are 95% confidence intervals.

## References

[1] Office for National Statistics. Prevalence of ongoing symptoms following coronavirus (COVID-19) infection in the UK: 2 February 2023. Available at: https://www.ons.gov.uk/peoplepopulationandcommunity/healthandsocialcare/conditionsanddiseases/bulletins/prevalenceofongoingsymptomsfollowingcoronaviruscovid19infectionintheuk/2february2023. Accessed 02 February 2023.

[2] Office for National Statistics. Coronavirus (COVID-19) Infection Survey, characteristics of people testing positive for COVID-19, UK: 16 November 2022. Available at: https://www.ons.gov.uk/peoplepopulationandcommunity/healthandsocialcare/conditionsanddiseases/bulletins/coronaviruscovid19infectionsurveycharacteristicsofpeopletestingpositiveforcovid19uk/16november2022. Accessed 01 February 2023.

[3] Long Covid Support and Long Covid Kids. How do Covid reinfections affect Long Covid? Results from an internet survey of people with Long Covid. August 2022. Available at: https://www.longcovid.org/images/Documents/Reinfections_in_Long_Covid_Survey_Report_by_Long_Covid_Support_and_Long_Covid_Kids_080922.pdf Accessed 01 February 2023.

[4] Bowe B, Xie Y, Al-Aly Z. Acute and postacute sequelae associated with SARS-CoV-2 reinfection. Nat Med, 2022; 28: 2398–405.

[5] UK Health Security Agency. NHS Test and Trace Statistics (England): methodology. 18 May 2022. Available at: https://www.gov.uk/government/publications/nhs-test-and-trace-statistics-england-methodology/nhs-test-and-trace-statistics-england-methodology. Accessed 01 February 2023.

[6] Pritchard E, Matthews PC, Stoesser N, et al. Impact of vaccination on new SARS-CoV-2 infections in the United Kingdom. Nat Med, 2021; 27: 1370–8.

[7] Office for National Statistics. Coronavirus (COVID-19) Infection Survey, characteristics of people testing positive for COVID-19, UK: 19 October 2022. Available at: https://www.ons.gov.uk/peoplepopulationandcommunity/healthandsocialcare/conditionsanddiseases/bulletins/coronaviruscovid19infectionsurveycharacteristicsofpeopletestingpositiveforcovid19uk/19october2022#measuring-the-data Accessed 01 February 2023.

[8] Davis HE, Assaf GS, McCorkell L, et al. Characterizing long COVID in an international cohort: 7 months of symptoms and their impact. EClinicalMedicine, 2021; 38: 101019.

[9] Michelen M, Manoharan L, Elkheir N, et al. Characterising long COVID: a living systematic review. BMJ Glob Health, 2021; 6: e005427.

[10] Ziauddeen N, Gurdasani D, O’Hara ME, et al. Characteristics and impact of Long Covid: Findings from an online survey. PLoS One, 2022; 17: e0264331.

[11] Austin PC. An introduction to propensity score methods for reducing the effects of confounding in observational studies. Multivariate Behav Res, 2011; 46: 399–424.

[12] UK Government. Coronavirus (COVID-19) in the UK. 2023. Available at: https://www.coronavirus.data.gov.uk Accessed 29 March 2023.

[13] Thompson EJ, Williams DM, Walker AJ, et al. Long COVID burden and risk factors in 10 UK longitudinal studies and electronic health records. Nat Commun, 2022; 13:3528.

[14] Ayoubkhani D, Bosworth ML, King S, et al. Risk of Long COVID in people infected with Severe Acute Respiratory Syndrome Coronavirus 2 after 2 doses of a Coronavirus Disease 2019 vaccine: community-based, matched cohort study. Open Forum Infect Dis, 2022; 9: ofac464.

[15] Antonelli M, Penfold RS, Merino J, et al. Risk factors and disease profile of post-vaccination SARS-CoV-2 infection in UK users of the COVID Symptom Study app: a prospective, community-based, nested, case-control study. Lancet Infect Dis, 2022; 22: 43–55.

[16] Watanabe A, Iwagami M, Yasuhara J, et al. Protective effect of COVID-19 vaccination against long COVID syndrome: A systematic review and meta-analysis. Vaccine, 2023; 41: 1783–90.

[17] Office for National Statistics. Self-reported long COVID after infection with the Omicron variant in the UK: 18 July 2022. Available at: https://www.ons.gov.uk/peoplepopulationandcommunity/healthandsocialcare/conditionsanddiseases/bulletins/selfreportedlongcovidafterinfectionwiththeomicronvariant/18july2022. Accessed 01 February 2023.

[18] Antonelli M, Pujol JC, Spector TD, et al. Risk of long COVID associated with delta versus omicron variants of SARS-CoV-2. Lancet, 2022; 399: 2263–64.

[19] COVID-19 Forecasting Team. Past SARS-CoV-2 infection protection against reinfection: a systematic review and meta-analysis. Lancet, 2023; 401: 833–42.

[20] Bobrovitz N, Ware H, Ma X, et al. Protective effectiveness of previous SARS-CoV-2 infection and hybrid immunity against the omicron variant and severe disease: a systematic review and meta-regression. Lancet Infect Dis, 2023; DOI: https://doi.org/10.1016/S1473-3099(22)00801-5

[21] Deng J, Ma Y, Liu Q, et al. Severity and outcomes of SARS-COV-2 reinfection compared with primary infection: a systematic review and meta-analysis. Int J Environ Res Public Health, 2023; 20: 3335.

[22] Iwasaki A, Putrino D. Why we need a deeper understanding of the pathophysiology of long COVID. Lancet Infect Dis, 2023; 23: 393–5.

[23] Pantelic M, Ziauddeen N, Boyes M, et al. Long Covid stigma: estimating burden and validating scale in a UK-based sample. PLoS ONE, 2022; 17: e0277317.

[24] Walker AJ, MacKenna B, Inglesby P, et al. Clinical coding of long COVID in English primary care: a federated analysis of 58 million patient records in situ using OpenSAFELY. Br J Gen Pract, 2021; 71: e806–14.

[25] Department of Health and Social Care. First UK cases of Omicron variant identified. 27 November 2021. Available at: https://www.gov.uk/government/news/first-uk-cases-of-omicron-variant-identified Accessed 07 March 2023.

